# Machine Learning for COVID-19 Patient Management: Predictive Analytics and Decision Support

**DOI:** 10.1101/2024.02.26.24303208

**Authors:** Christopher El Hadi, Rindala Saliba, Georges Maalouly, Moussa Riachy, Ghassan Sleilaty

## Abstract

**Background:** The global impact of severe acute respiratory syndrome coronavirus 2 (SARS-CoV-2) has profoundly affected economies and healthcare systems around the world, including Lebanon. While numerous meta-analyses have explored the systemic manifestations of COVID-19, few have linked them to patient history. Our study aims to fill this gap by using cluster analysis to identify distinct clinical patterns among patients, which could aid prognosis and guide tailored treatments.

**Methods:** We conducted a retrospective cohort study at Beirut’s largest teaching hospital on 556 patients with SARS-CoV-2. We performed cluster analyses using K-prototypes, KAMILA and LCM algorithms based on 26 variables, including laboratory results, demographics and imaging findings. Silhouette scores, concordance index and signature variables helped determine the optimal number of clusters. Subsequent comparisons and regression analyses assessed survival rates and treatment efficacy according to clusters.

**Results:** Our analysis revealed three distinct clusters: “resilient recoverees” with varying disease severity and low mortality rates, “vulnerable veterans” with severe to critical disease and high mortality rates, and “paradoxical patients” with a late presentation but eventual recovery.

**Conclusions:** These clusters offer insights for prognosis and treatment selection. Future studies should include vaccination data and various COVID-19 strains for a comprehensive understanding of the disease’s dynamics.

## 1. Background

Over the past three years, the epidemic of severe acute respiratory syndrome caused by the severe acute respiratory syndrome coronavirus 2 (SARS-CoV-2) has spread around the world, affecting multiple economies and healthcare systems, and rubbing salt into the wound of Lebanon’s economy and status quo [1]. As an infectious disease of the respiratory tract, SARS-CoV-2, which causes coronavirus disease (COVID-19), generally manifests itself with common symptoms such as fever, fatigue, headache, cough and sore throat [2]. However, clinical presentations and disease severity in patients with COVID-19 can vary considerably, depending on the circulating viral strain, comorbidities and the patient’s immune constitution. Symptoms can range from none, as in over a third of infected individuals, to the life-threatening, as in those with underlying acute respiratory failure [3]. Predicting a patient’s reaction to the virus and administering the appropriate treatments to avoid an unfavorable and potentially fatal outcome may seem an avant-garde approach, but it’s possible thanks to modern statistical algorithms for clustering patients.

Cluster analysis is a fundamental technique in data mining, designed to reveal patterns that may be hidden by the complexity of the data, and extract knowledge from them. It has many applications in a variety of fields, e.g. socio-economic and medical, and has proved particularly useful for uncovering patterns in clinical data that might not be easily discerned by human analysis alone [4,5]. It involves the use of algorithms to divide data into groups of observations or “clusters”, based on increasing the similarity between the components of a cluster, while reinforcing the dissimilarity between clusters [6]. Such advance have led to a paradigm shift in medicine, where precision medicine is becoming tantamount to evidence-based medicine, particularly in areas such as cancer and metabolic diseases [7]. For challenging diseases such as COVID-19, it may be of interest to identify distinct patient categories to enable a more personalized and rigorous approach to patient care.

We thus sought to classify patients with COVID-19 treated at the Hôtel Dieu de France Hospital in Beirut, on the basis of their medical history and the biochemical and radiological results obtained on hospital presentation. The representative criteria of each class obtained were then compared, enabling the clusters to be calibrated in order to adopt proactive approaches for each Lebanese admitted patient newly diagnosed with COVID-19. The latter will have the opportunity to be matched with one of the studied clusters of COVID-19 patients with clear, albeit probabilistic, treatment recommendations ready for implementation.

## 2. Methods

We conducted a single-center retrospective cohort study. We included 556 hospitalized patients with confirmed COVID-19 between September 22, 2019, and October 12, 2021. All statistical analyses were performed using R 4.3.1 (The R Foundation for Statistical Computing, Vienna, Austria) [8].

### 2.1. Data collection

The data were extracted from the Hôtel Dieu de France hospital electronic database. Only the first data values collected within 24 hours of hospital admission were used for cluster analysis (Table 1). The prognostic value of the different treatments administered was assessed by studying their effect on severity variables such as contraction of nosocomial infections, development of pneumo-mediastinum, composite fatal outcome (i.e admission, intubation and death), day of ICU transfer, date of intubation, occurrence of thromboembolic or hemorrhagic events and the corresponding dates, duration of hospitalization and all-cause death.

**Table 1.**
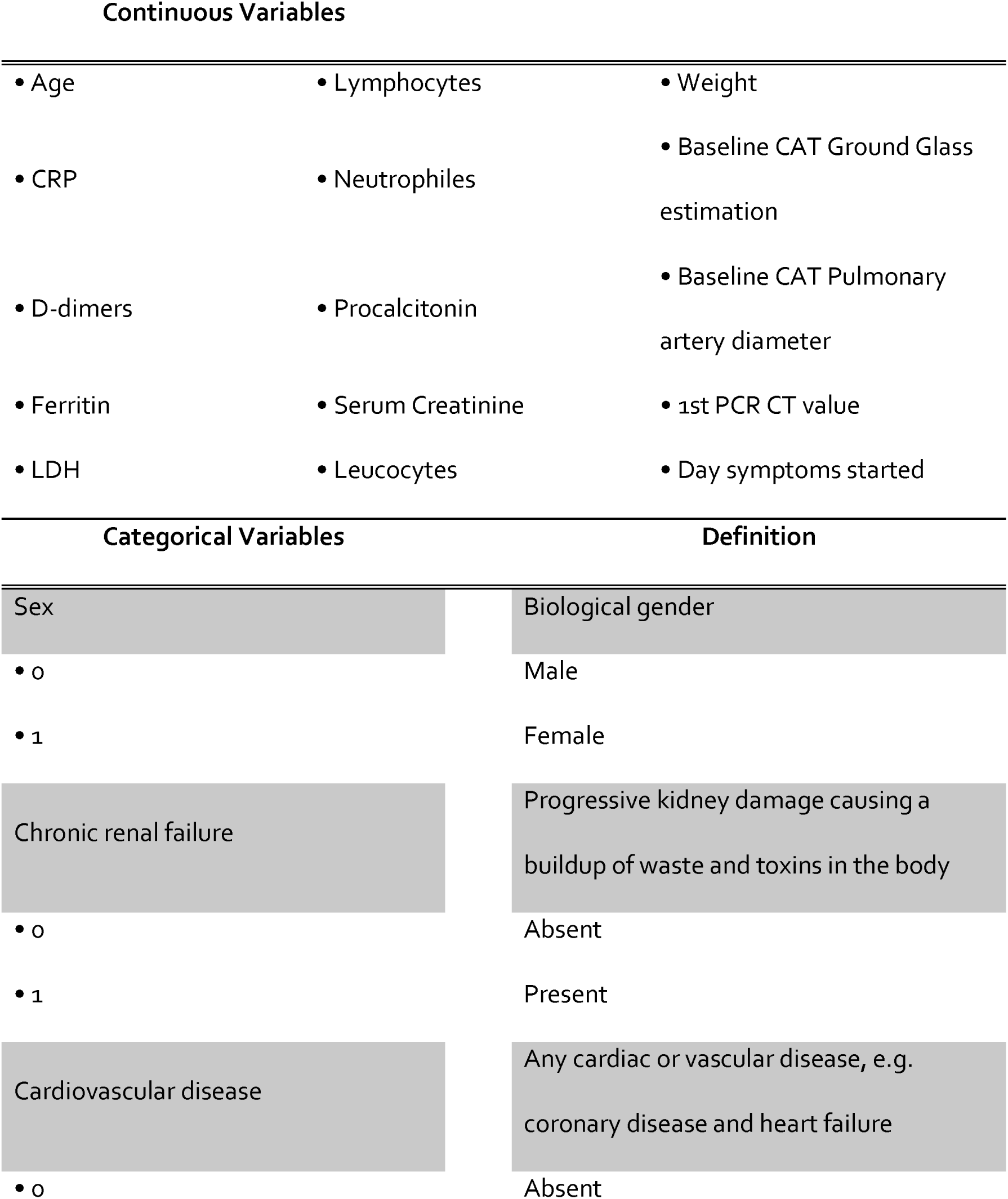
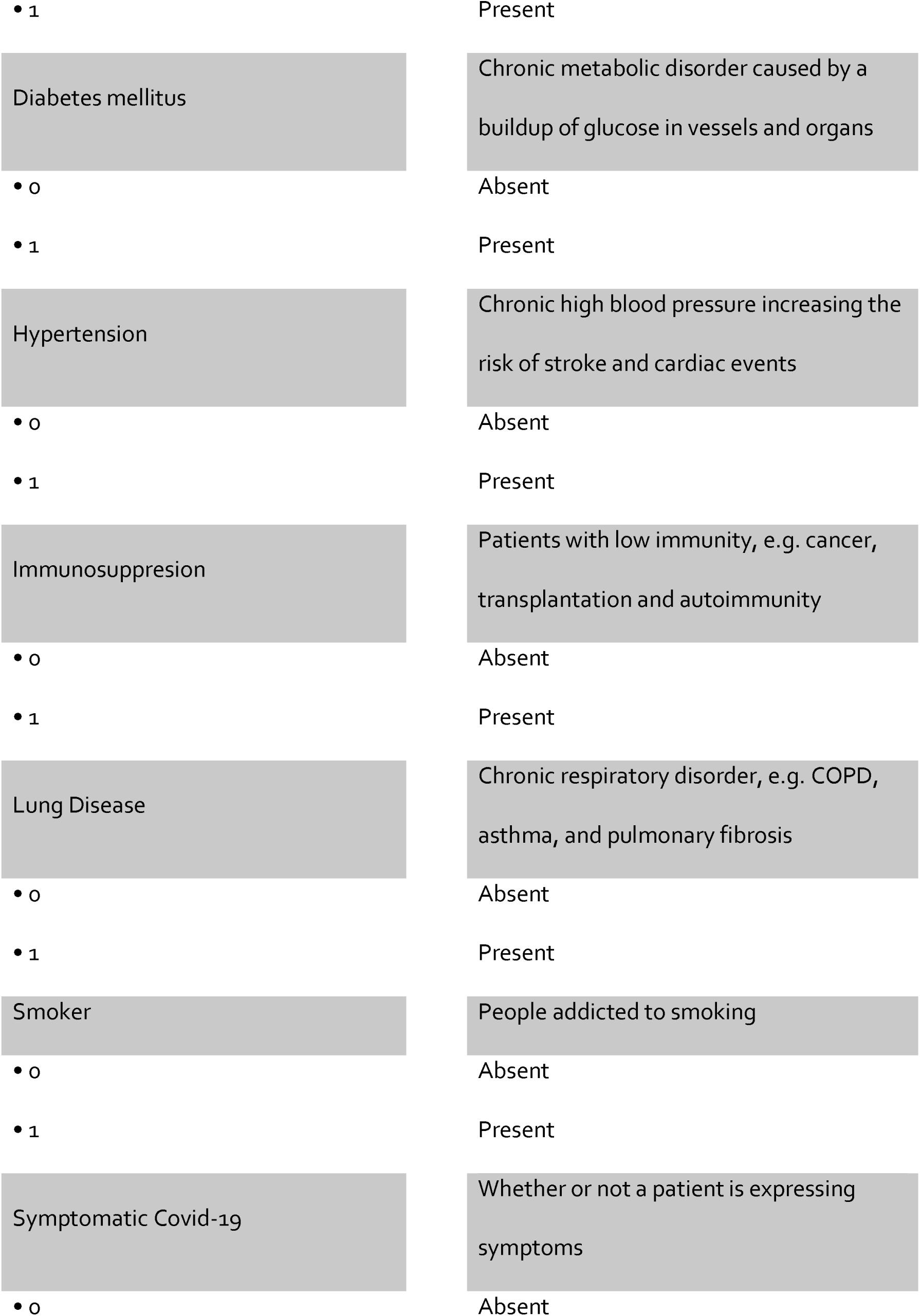
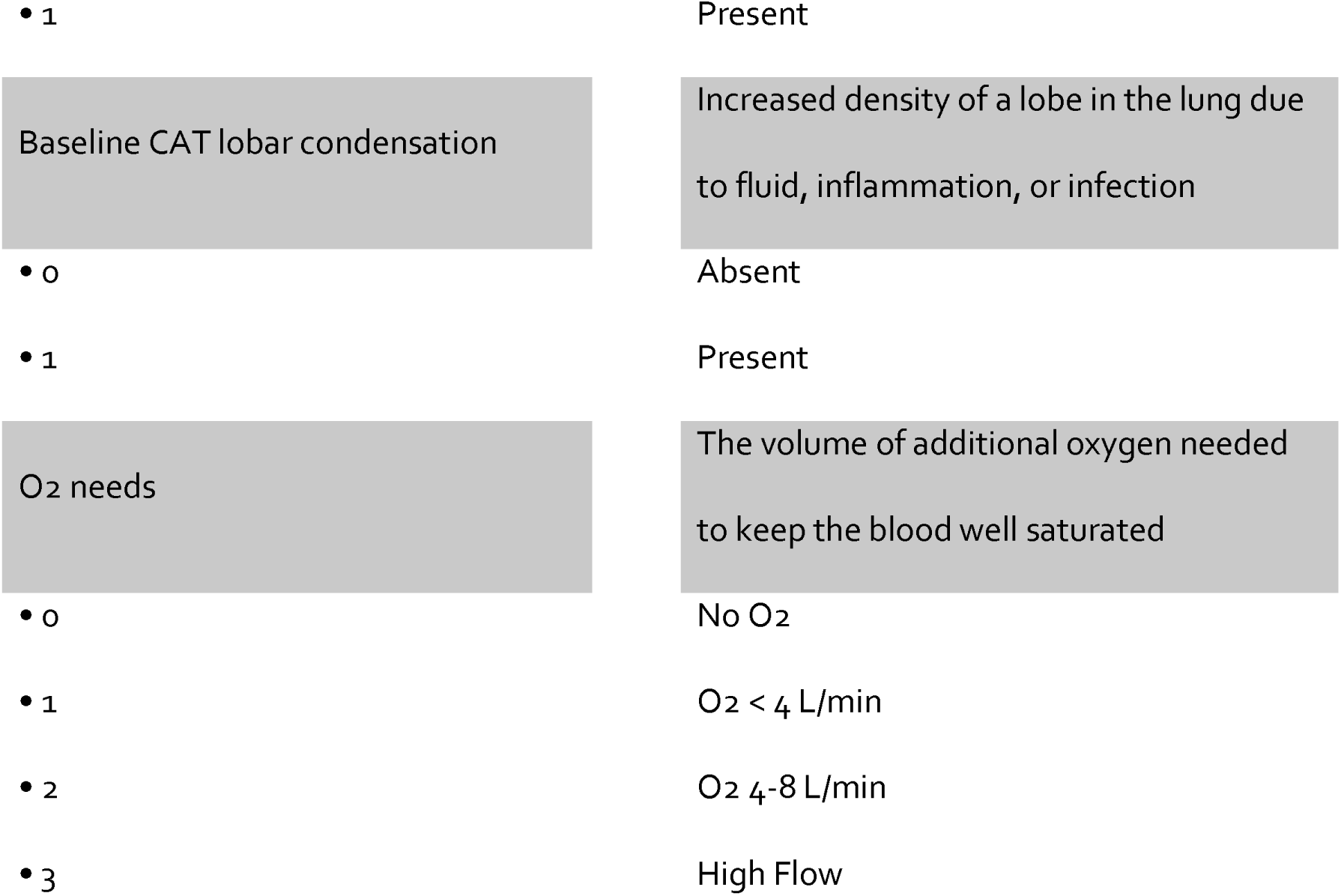
Parameters used for patient clustering. The values used in the algorithm for categorical parameters are illustrated.

### 2.2. Data pre-processing and Clustering

The data studied is composed of continuous and categorical/ordinal variables (Table 1). Scaling and normalization were applied to continuous variables to meet the requirements of each algorithm [9,10]. Missing data were handled using the *mice* package [11].

### 2.3. Cluster Analysis

The study used the 26 variables described in Table 1 and three clustering methods (K-prototypes, KAMILA [12] and LCM [13]) to cluster COVID-19 patients. The appropriate number of clusters was determined using silhouette scores [14] and Harrel’s concordance index [15]. The clinical relevance of the results was assessed on the basis of differences in survival and hospital stay between clusters, and the identification of signature variables likely to differentiate clusters. Further information on the clustering process can be found in Supplementary file 1.

### 2.4. Statistical analysis

Kaplan-Meier risk curves, logistic and Cox regressions were applied to 4 subclusters of patients, each defined by whether they were admitted to the ICU and whether they were superinfected or not (procalcitonin ≥0.5). Odd ratio (OR) and Hazard Ratio (HR) with 95% Confidence Interval (CI) were used to assess treatment effects. Mann-Whitney U or Kruskal-Wallis tests were used for continuous variables, followed by Dunn’s post-test if the latter test was used [16]. The Chi-2 test (and Goodness-of-Fit test) and Fisher’s exact test were used to compare categorical variables, followed by a post hoc test using Bonferroni correction [17].

## 3. Results

### 3.1. Comparison of characteristics

After performing a systematic and comparative analysis of the three algorithms considered, namely K-prototypes, KAMILA and LCM, KAMILA proved to be the best clustering algorithm because it had the highest silhouette score and C-index, and produced the highest number of signature features, indicating superior clustering quality and differentiation ability compared to the other methods. A comprehensive, meticulously-documented, step-by-step analysis is made available for reference in Supplementary File 1.

Table 2 summarizes the demographic data and days since symptom onset of our sample, while Table 3 summarizes that of each cluster. Cluster 1 was the largest (239 patients) in contrast to the 2^nd^ and 3^rd^ clusters, both having almost the same number of patients (153 and 156, p = 0.865 for pairs 2-3). Roughly, Cluster 1 had the youngest patients with 5 out of 6 patients being ≤ 68 years, Cluster 2 had the oldest patients with the same ratio being ≥ 69 years, and Cluster 3 had the older half of Cluster 1 and the younger half of Cluster 2. Cluster 1 had the lowest weighted patients with Cluster 3 consisting mostly of the heavier. Cluster 1 had an almost equal distribution of genders, unlike Cluster 2 and 3 having predominantly men. Cluster 2 patients presented the earliest to the hospital, whereas for Cluster 3 the latest.

**Table 2.**
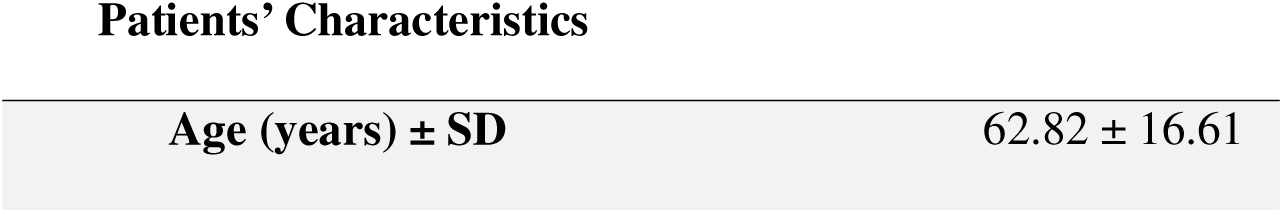
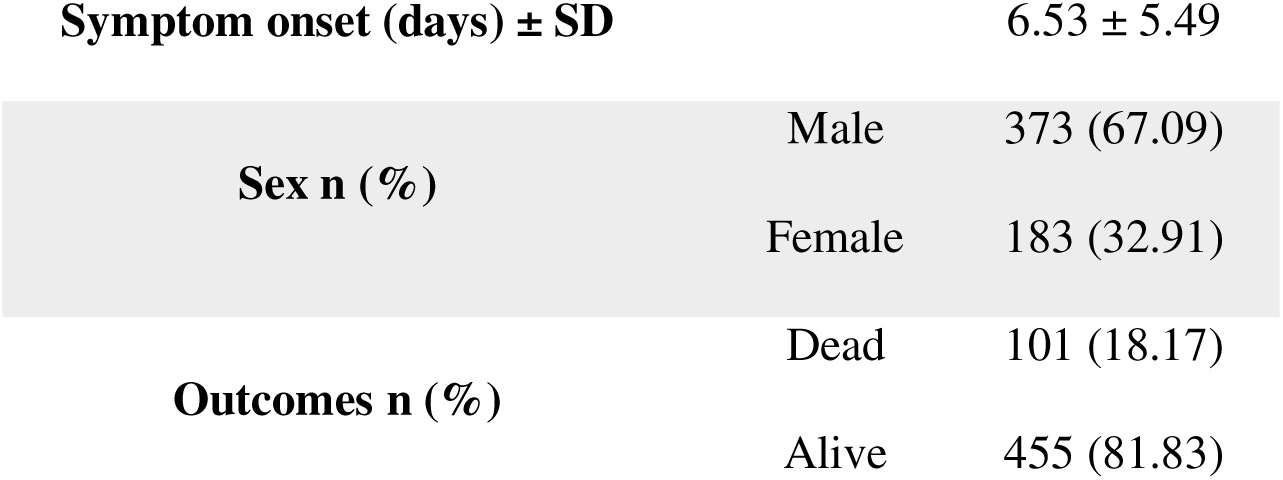
Demography and days since symptom onset of 556 COVID-19 patients.

Patients' Characteristics
|  |  |
| --- | --- |
| Age (years) ± SD | 62.82 ± 16.61 |

|  |  |  |
| --- | --- | --- |
| **Symptom onset (days) ± SD** |  | 6.53 ± 5.49 |
| **Sex n (%)** | Male | 373 (67.09) |
|  | Female | 183 (32.91) |
| **Outcomes n (%)** | Dead | 101 (18.17) |
|  | Alive | 455 (81.83) |

**Table 3.**
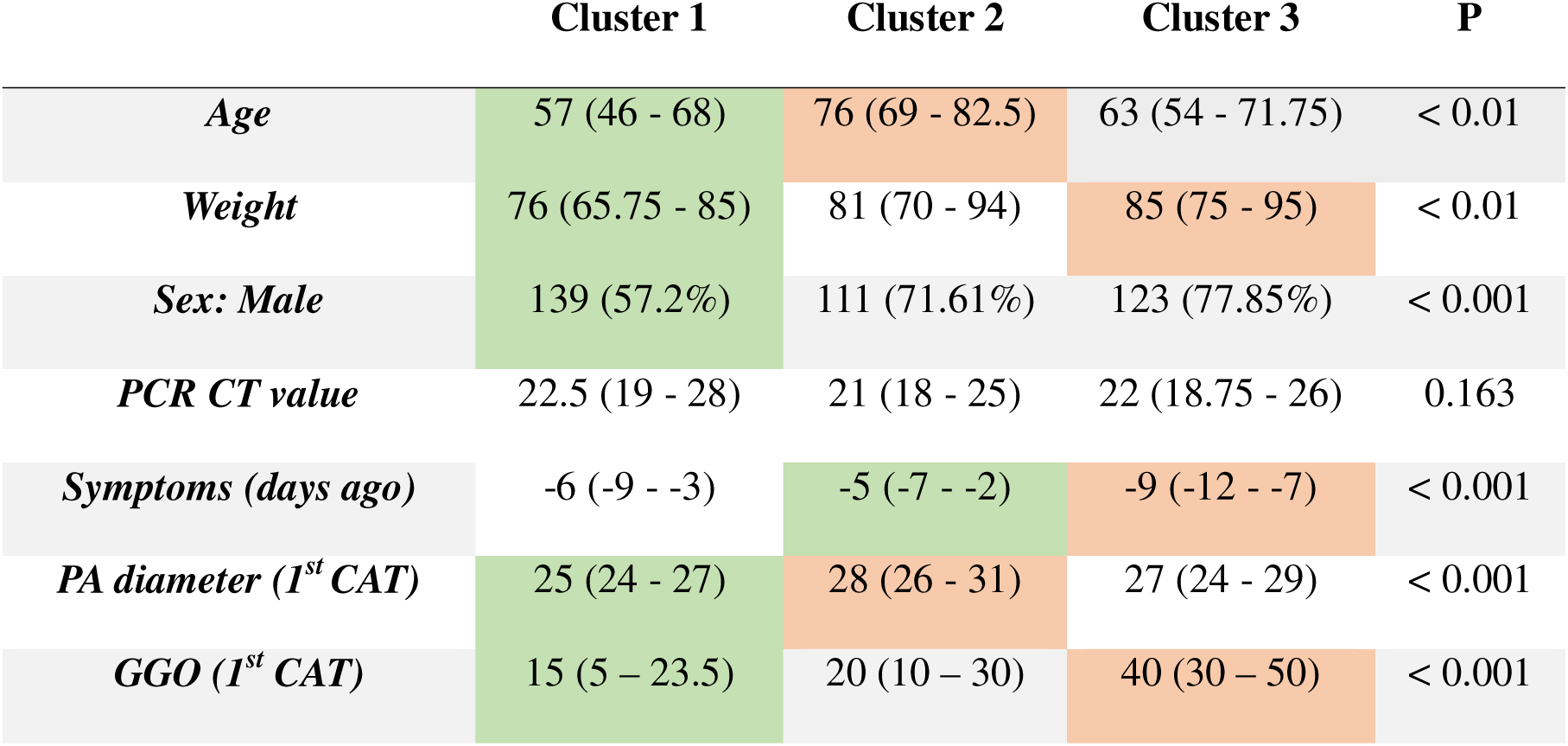
Anthropometric and demographic measurements of three COVID-19 clusters identified using KAMILA cluster analysis. Grey cells in a row: 1 = all pairs significantly different. Green = lowest, Orange = highest. Abbreviations: PA pulmonary artery, CAT computerized axial tomography, GGO: ground-glass opacities

|  | **Cluster 1** | **Cluster 2** | **Cluster 3** | **P** |
| --- | --- | --- | --- | --- |
| *Age* | 57 (46 - 68) | 76 (69 - 82.5) | 63 (54 - 71.75) | < 0.01 |
| *Weight* | 76 (65.75 - 85) | 81 (70 - 94) | 85 (75 - 95) | < 0.01 |
| *Sex: Male* | 139 (57.2%) | 111 (71.61%) | 123 (77.85%) | < 0.001 |
| *PCR CT value* | 22.5 (19 - 28) | 21 (18 - 25) | 22 (18.75 - 26) | 0.163 |
| *Symptoms (days ago)* | -6 (-9 - -3) | -5 (-7 - -2) | -9 (-12 - -7) | < 0.001 |
| *PA diameter (1<sup>st</sup> CAT)* | 25 (24 - 27) | 28 (26 - 31) | 27 (24 - 29) | < 0.001 |
| *GGO (1<sup>st</sup> CAT)* | 15 (5 - 23.5) | 20 (10 - 30) | 40 (30 - 50) | < 0.001 |

Cluster 1 has the fewest hypertensive and diabetic patients, but percentages comparable to Cluster 3 for other risk factors, while Cluster 2 has the highest rates for all risk factors. Roughly, half of Cluster 1 did not require any oxygen, unlike half of Cluster 3 needing higher flow of oxygen. Cluster 1 had the lowest laboratory values including IL6, except for lymphocyte count, LDH and Ferritin being comparable to Cluster 2. While the latter exhibited the highest procalcitonin and creatinine levels, Cluster 3 had the highest leucocyte (viz. neutrophile) count, CRP, LDH and Ferritin but lowest Lymphocyte count. Cluster 3 had the most ground-glass opacities on serial CT scans while Cluster 2 had the largest pulmonary artery diameter. Lastly, Cluster 1 has the lowest rates of ICU admissions and intubations, Cluster 2 the highest rates of hemorrhagic events and Cluster 3 the highest rates of thromboembolic events. Relevant patient risk factors are shown in Table 4, lab results in Table 5 and hospital outcomes in Table 6.

**Table 4.** Relevant risk factors in the three COVID-19 clusters obtained using KAMILA. Grey cells in a row: 1 = all pairs significantly different, 2 = those aren’t significantly different. † = slightly different. Green = lowest, Orange = highest.

| Risk Factors | Cluster 1 | Cluster 2 | Cluster 3 | P |
| --- | --- | --- | --- | --- |
| ***Hypertension*** | 78 (32.1%) | 147 (94.84%) | 79 (50%) | < 0.001 |
| ***Diabetes*** | 28 (11.52%) | 86 (55.48%) | 43 (27.22%) | < 0.001 |
| ***Coronary Artery Disease*** | 24 (9.88%) | 91 (58.71%) | 26 (16.46%) | < 0.001 |
| ***Pulmonary Diseases*** | 32 (13.79%) | 49 (32.03%) | 23 (14.94%) | < 0.001 |
| ***Heart Failure*** | 1 (4.55%) | 13 (30.95%)† | 4 (7.14%) | 0.002 |
| ***Chronic Kidney Disease*** | 12 (4.94%) | 66 (42.58%) | 5 (3.16%) | < 0.001 |
| ***Smoking*** | 34 (14.66%) | 56 (36.6%) | 38 (24.68%) | < 0.001 |
| ***Immunocompromised*** | 22 (9.05%) | 18 (11.61%) | 10 (6.33%) | 0.272 |

**Table 5.** Laboratory findings in the three distinct COVID-19 clusters. Number of grey cells in a row: 1 = all pair-wise comparisons are significant, 2 = this comparison isn’t significant. † = slightly different. Green = lowest, Orange = highest.

|  | Cluster 1 | Cluster 2 | Cluster 3 | P |
| --- | --- | --- | --- | --- |
| ***Leukocytes*** | 5850<br>(4525 - 7700) | 7300<br>(5300 - 9200) | 9700<br>(7000 - 12550) | < 0.001 |
| ***Serum creatinine*** | 66<br>(53 - 80) | 121<br>(88 - 201) | 74<br>(62 - 93) | < 0.001 |
| ***Procalcitonin*** | 0.11<br>(0.06 - 0.22) | 0.32<br>(0.14 - 0.94) | 0.195<br>(0.1 - 0.4) | < 0.001 |
| ***Neutrophils*** | 4170<br>(3060 - 5940) | 5380<br>(3875 - 7505) | 8105<br>(5678 - 11010) | < 0.001 |
| ***Lymphocytes*** | 1015<br>(630 - 1440) | 830<br>(580 - 1265) | 690<br>(483 - 1025) | < 0.001 |
| ***LDH*** | 275.5<br>(219 - 349) | 291.5<br>(235 - 366) | 488.5<br>(394 - 649) | < 0.001 |
| ***Ferritin*** | 611<br>(277 - 1078) | 579<br>(286 - 1044) | 1196<br>(674 - 2152) | < 0.001 |
| ***D-Dimer*** | 0.58<br>(0.36 - 1.23) | 1.23<br>(0.63 - 2.77) | 0.995<br>(0.59 - 1.71) | < 0.001 |
| ***CRP*** | 49<br>(19 - 93) | 94.25<br>(46 - 169) | 136.5<br>(79 - 189.5) | < 0.001 |
| ***IL6*** | 24.6<br>(13.3 - 94) | 41<br>(29.7 - 97.5) | 47.35<br>(19.25 - 117.3) | 0.071 |

**Table 6.**
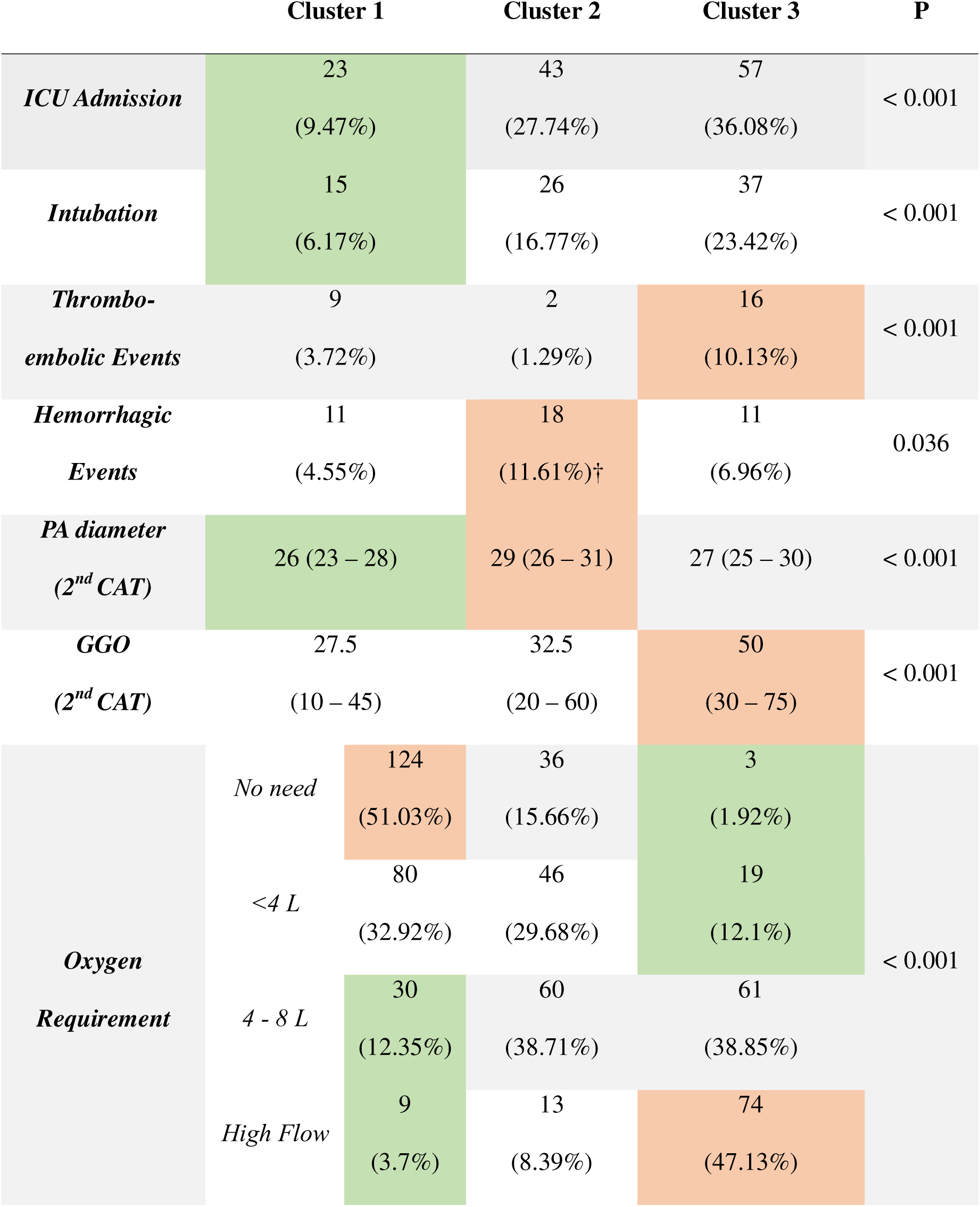
Outcomes during hospitalization of the three distinct COVID-19 clusters. Number of grey cells in a row: 1 = all pair-wise comparisons are significant, 2 = this comparison isn’t significant. † = slightly different. Green = lowest, Orange = highest. Abbreviations: PA pulmonary artery, CAT computerized axial tomography, GGO: ground-glass opacities.

### 3.2. Survival analysis

Survival analysis is a statistical method used to study the time until an event of interest occurs, like patient deaths. In the classical approach, it examines the probability of an event occurring: an HR > 1 indicates an increased probability of the event occurring and HR < 1 indicates a decreased probability. However, a modified analysis will be featured here, where deaths are considered discharges, hence HR > 1 will indicate a positive outcome, i.e. faster discharge.

The analysis suggests that cluster 2 had the highest risk of all-cause death, while Clusters 1 and 3 were not different in terms of mortality. Moreover, clusters 2 and 3 have significantly higher risks of prolonged stay resulting in faster Cluster 1 patient discharges. The results of the two analyses are summarized in Table 7.

**Table 7.** Overview of key outcomes for the three COVID-19 clusters. Durations are represented as medians in days. † HR of discharge < 1 is counter intuitively a bad outcome.

| Cluster | N | Mortality | Survival<br>Time | HR<br>Death | Length<br>of Stay | HR<br>Discharge† |
| --- | --- | --- | --- | --- | --- | --- |
| 1 | 239 | 7.11% | 49 | Reference | 6 | Reference |
| 2 | 153 | 34% | 41 | 2.68<br>(1.54-4.66) | 11 | 0.51<br>(0.40-0.65) |
| 3 | 156 | 13.4% | 41 | 1.26<br>(0.70-2.29) | 11 | 0.53<br>(0.42-0.66) |

Based on all the previous results, the clusters will be hereafter labeled according to the population they describe. Namely, Cluster 1 will be dubbed “Resilient Recoverees”, Cluster 2 “Vulnerable Veterans,” and Cluster 3 “Paradoxical Patients”.

### 3.3. Regression analysis

The association of various treatments with multiple outcomes was evaluated within each cluster to minimize complications and unnecessary interventions. Detailed treatment results, including HR and OR, are available in Supplementary file 2. Subclusters were constructed based on whether PCT at admission ≥ 0.5 and admission location (ICU or regular wards, as shown in Table 7). Mean and SD for treatment initiation, duration, and doses is presented in Table A (Supplementary file 2).

#### Non-superinfected non-ICU resilient recoverees

The use of carbapenem treatment was associated with longer hospital stays. In contrast, Tocilizumab doses of ~750 mg or more were associated with faster patient discharge.

#### Superinfected non-ICU resilient recoverees

Aminoglycosides, glucocorticoid treatment equal to or greater than ~6 weeks, and azithromycin at ~1.5 weeks from symptom onset or later were correlated with prolonged hospitalization periods and provided no benefit.

#### Superinfected vulnerable veterans

Non-ICU patients required oxygen therapy ranging from 4 to 8 liters which extended their hospital stays. The odds of reaching the composite outcome (ICU admission, intubation, then death) if a patient duration of glucocorticoid therapy averaged ~3 weeks and they did not receive aminoglycoside treatment averaged 0.220 (0.054 – 0.619).

#### Non-superinfected ICU vulnerable veterans

Deferred ICU admissions were associated with a delay of ~1.5 weeks in starting Tocilizumab and/or glucocorticoid therapy from onset of symptoms. This also applied to Hydroxychloroquine treatment if it extended beyond 3 weeks. This suggests that all 3 therapies were started after ICU admission, not preventively before. Conversely, early ICU admissions were associated with the administration of doses of Aspirin ≥ 324mg. Regardless administration date, the mentioned therapies did not benefit the patient.

#### Non-superinfected non-ICU vulnerable veterans

Cephalosporins failed to impact the composite outcome, proving its uselessness. Antibiotic therapy beyond ~2 weeks benefitted patients suggested later superinfection. Glucocorticoid use for ~6 weeks or more offered no advantage other than prolonging stays. Similarly, glycopeptides extended hospital stays and were associated with increased bleeding risk, suggesting their use as poor prognostic indicator.

#### Non-superinfected non-ICU paradoxical patients

An increase in mortality was significantly linked to the use of prophylactic doses of antiplatelets in comparison to alternative dosage regimens of the same drug. The administration of doxycycline and the implementation of prone positioning were associated with expedited discharges, as opposed to those subjected to glucocorticoid treatment for a duration of ~6 weeks or more and prednisone exceeding 25 mg.

#### Non-superinfected ICU paradoxical patients

Glucocorticoid therapies that lasted beyond ~6 weeks also delayed ICU admission. Delayed ICU admission was associated with very early and multiple courses of Tocilizumab or Baricitinib treatment lasting beyond ~2 weeks, but there was no advantage in starting Tocilizumab thereafter. Subsequent superinfections and abstaining from using Tocilizumab were the main promotors of ICU admission. As for antibiotics, expecting to administer glycopeptides or cotrimoxazole at ~2 weeks from symptom onset if a bacterium was elucidated could delay ICU admissions. During ICU stay, neither carbapenems nor azithromycin had a positive impact on patient survival. The use of Remdesivir did not show any benefit.

## 4. Discussion

Over the past three years, SARS-COV-2 has spread globally manifesting itself in different clinical presentations ranging from a fleeting flu to critical illness [18]. Few studies have linked pathophysiologic findings to identify patient patterns for personalized treatments [19,20]. Cluster analysis is a promising approach to categorizing patients, and our study classified patients based on medical history, biochemistry, and radiology. KAMILA algorithm produced the best results [12], identifying three distinct patient clusters with unique characteristics. This approach challenges the most updated treatment guidelines that rely on univariate patient data that has not been interpreted using a multivariate approach, such as clustering. Specifically, it challenges the COVID-19 clinical spectrum outlined in the frequently updated NIH treatment guidelines [18]. As an overview, adults were considered to have asymptomatic or pre-symptomatic infection; mild, symptomatic illness without stigmata of pneumonia; moderate illness, with signs of lower respiratory tract disease but SpO2 ≥94% on room air; severe illness, including patients with SpO2 <94% on room air who are not in shock or respiratory and organ failure; and critical illness with system failure. This classification is primarily based on oxygen saturation, a measurement that can be inconclusive, particularly in people aged ≥50 years with high-risk comorbidities and severe outcomes. NIH recommendations on oximetry interpretation favor consideration of the patient’s overall clinical presentation and history. That said, it is essential to be proactive and detect and therefore treat those having risky histories so as to avoid admissions to the IC, invasive mechanical ventilation, and death. CDC researchers reviewed the risk factors that favor COVID-19 progression into severe statuses [21], which have been taken into account in the therapeutic management of hospitalized patients as per the NIH recommendations [22]. Our study aims to refine these predictive criteria through clustering.

To illustrate, the resilient recoverees, the largest cluster having the fastest recovery and lowest mortality, can be considered a moderate-to-severe COVID-19 group. They include middle aged fit patients, with markedly few cardiovascular risk factors, e.g. hypertension. They can benefit from minimal (e.g. < 8 L O2) to no therapeutic approaches considering the low mortality burden. Laboratory results were roughly normal with no markers of severe COVID-19 [23]. It’s worth noting that IL-6 levels in this cluster were the lowest, but the lack of significant difference from other clusters is unreliable due to the infrequency of IL-6 measurements. The use of Tocilizumab may accelerate the discharge of non-ICU patients with no bacterial superinfections, but its cost-effectiveness and priority in this cluster should be minded.

As for the vulnerable veterans, they mostly include elderly men who have multiple risk factors and multiple comorbidities. They are at higher risk of severe to critical COVID-19 with more hemorrhagic events, superinfections and a higher mortality rate, most likely favored by their comorbidities. In addition, these patients had the largest pulmonary artery and the highest serum creatinine and procalcitonin suggesting the presence of pulmonary hypertension and the prevalence of superinfections. That said, superinfected ICU patients appeared to benefit from a ~ 3 week course of glucocorticoid therapy, which aligns with the NIH recommendations [18].

The paradoxical patient cluster, predominantly middle to old age men with nearly twice commoner hypertensive and diabetic patients than resilient recoverees, presenting 1 to 2 weeks late with more thromboembolic events. Despite severe to critical COVID-19 classification by NIH guidelines, their mortality rates mirror resilient recoverees. Paradoxically, if one were to consider merely their history, they would risk misclassifying the cluster as resilient. This cluster experiences prolonged stays akin to vulnerable veterans, similarly demanding ICU admissions and intubations, but requiring noticeably greater high-flow O2 supplementation. Glass opacities on CT scans predominate and are associated with elevated COVID-19 severity biomarkers [23], notably lymphopenia but also CRP, ferritin and LDH alluding to a “cytokine storm”. This excessive, uncontrolled response contribute to tissue damage, organ failure, and heightened mortality [24]. Ancillary should aid in distinguishing resilient from paradoxical patients, emphasizing the impact of the virus itself. For non-superinfected regular wards patients, recent literature echoed our conclusions on the effectiveness of prone positioning [18] and doxycycline therapy [25], particularly in hastening discharge. More, it agrees on the uselessness of anti-platelet therapy in this group [18]. As for non-infected ICU patients, Tocilizumab or Baricitinib early on and for long periods delay ICU admissions, as well as glucocorticoid therapy for more than 6 weeks.

Two studies have also attempted to cluster COVID-19 patients into groups. The first study by Han *et al.* used factor analysis for mixed data (FAMD) [26]. What differentiates this study from ours is the addition of patient-experienced symptoms. Their results showed that the patients could be divided into three distinct clusters: Cluster A, the most severe with the longest hospital stays; Cluster B, of intermediate severity COVID-19 with a length of stay as long as Cluster A; and Cluster C, the mildest with the shortest length of stay. Their analysis showed that cluster A had the worst survival rate, whereas cluster B had higher CRP, D-dimer, AST, and LDH levels, indicating a quintessential COVID-19 phenotype. Clusters A and B are thus comparable to our vulnerable and paradoxical patients, respectively. Cluster C had mainly systemic and digestive symptoms and a low frequency of typical symptoms of fever and cough; because of its low severity, it mostly resembles the resilient recoverees. Our study, in comparison, included imaging studies. We were also able to find significant correlation to age. Be that as it may, old age was proven to be associated with adverse outcomes for patients with COVID-19 [27].

Arévalo-Lorido *et al.,* the second study similar to ours, analyzed datasets by applying the Random Forest model and the Gaussian mixed model by clustering [28]. The algorithms generated six clusters, the last three of which had high mortality rates from any cause or ended up in intensive care, whereas the first three included patients who did not. The most important comorbidities were heart failure, atrial fibrillation, vascular disease, and neurodegenerative disease, which were mainly present in the last three clusters. The fifth cluster, with the poorest prognosis, included those with liver, kidney, and gastrointestinal diseases, as well as chronic obstructive pulmonary disease. From what has been described, the first three clusters converge on the resilient cluster, and the last three clusters converge on the vulnerable and paradoxical clusters, with cluster 5 in the study being the closest to the paradoxical patients. Contrasted to our study, it did not include data on imaging. Furthermore, KAMILA concisely separated patients into a small number of meaningful clusters, unlike Random Forest and Gaussian mixed model, which resulted in multiple clusters which seem unfathomable.

Our study has significant strengths. Specifically, we recognize the effectiveness of model-based algorithms in clustering mixed data, providing a rationale for this choice. Additionally, we incorporated imaging findings and pinpointed vulnerable age groups, all within an optimal small number of clusters. Finally, the article extends its analysis by investigating the impact of various treatments on four subtypes of patients within each cluster.

We acknowledge our study has some limitations. Firstly, the number of patients decreased significantly due to multiple stratifications, so further analysis with larger populations is recommended. Secondly, patients’ symptoms were not taken into account during data collection, which could have made the classification more clinically friendly. Thirdly, the PCT threshold used to consider patients as infected could have been improved by doing serial measures not only for PCT but also other markers in conjunction (e.g. CRP and imaging). Finally, the data collection was performed before vaccination campaigns and when one COVID-19 variant dominated the cases, so it would be interesting to study the effects of vaccination on the classes and the effect of different variants on patients to find a common classification for all COVID-19 strains.

## Data Availability

All data produced in the present study are available upon reasonable request to the authors

## Acknowledgments

We want to acknowledge the support of our institution and the interests of the contributors who have invested their time and expertise in this project. Moreover, this research has not received any external financial assistance or grants.

## Ethical Approval

The present study was conducted in accordance with the principles outlined in the Declaration of Helsinki. Ethical oversight for this study, specifically on the sole use of anonymized patient data, was obtained from the Institutional Review Board at Saint Joseph University affiliated hospital, Hotel-Dieu de France, in Beirut, Lebanon.

## Author contribution

C.H. drafted the manuscript and analyzed the data collected and supplied by M.R. and G.S.

G.M. supervised the work and analyzed the outputs. R.S. extensively revised all versions of the article and contributed to the final manuscript.

## Abbreviations

ASMD: Absolute standardized mean difference
ARDS: Acute respiratory distress syndrome
BIC: Bayesian information criterion
CI: Confidence interval
COVID-19: Coronavirus disease 2019
CRN: Creatinine
CRP: C-reactive protein
CT: Computed tomography
CT-PCR: Cyclic value - polymerase chain reaction
HR: Hazard ratio
ICU: Intensive care unit
IL-6: Interleukin-6
KAMILA: K-means of Mixed Large data
k: Number of clusters
LCM: Latent Class Models
LDH: Lactate dehydrogenase
OR: Odds ratio
PCT: Procalcitonin
PMM: Predictive mean matching
SARS-CoV-2: severe acute respiratory syndrome coronavirus 2
VIF: Variance inflation factor
WHO: World Health Organization

## Notes

### Competing Interest Statement

The authors have declared no competing interest.

### Funding Statement

This study did not receive any funding

### Author Declarations

We, the authors, obtained ethical approval for our study from the Ethics Committee of the Hospital Hotel-Dieu de France (HDF), Beirut, Lebanon, affiliated to the Faculty of Medicine of the Universite Saint Joseph (USJ), Beirut, Lebanon. The Ethics Committee is chaired by Dr. Sami Richa, psychiatrist; Prof. Michel Scheuer, Director of the University Center for Ethics at the USJ secretary; Prof. Georges Abi Tayeh, gynecologist at HDF; Prof. Nasri Antoine Diab, lawyer at the Beirut and Paris Bar Association; Mrs. Soha Abdel Malak, nurse at the HDF - expert in hospital hygiene; Dr. Jad Habib, Family Medicine, at the Centre universitaire de sante familiale et communautaire at USJ; Ms. Hyam Kahi, Social Assistant; Prof. Ronald Moussa, Neurosurgeon at HDF; Dr. May Fakhoury, Chief Pharmacist at HDF; Dr. Lina Iskandar Hawat, Dean's Representative at the Faculty of Religious Sciences at USJ; Mr. Ayad Wakim, Journalist; Ass. Prof. Eliane Nasser Ayoub, Anesthesiology and Resuscitation at HDF. The committee authorized the use of patient data from the Hotel-Dieu de France dataset on condition of anonymity.

