## Supplementary File 2 for "Machine Learning for COVID-19 Patient Management: Predictive Analytics and Decision Support"

**Variable Information**

| Variable | Position | Label | Domain | Measurement Level | |
|---|---|---|---|---|---|
| AGE | Scale | 2 | Age | IDENTIFICATION | |
| CL_O2_NEED | Ordinal | 3 | O2 needs | CLINICAL | |
| GENDER | Nominal | 4 | Gender | TREATMENT | |
| LB_CRP | Scale | 5 | CRP | BIOLOGY | |
| LB_D_DIMERS | Scale | 6 | D-dimers | BIOLOGY | |
| LB_FERRITIN | Scale | 7 | Ferritine | FILTERS FOR PROJECTS | |
| LB_LDH | Scale | 8 | LDH | BIOLOGY | |
| LB_LYMPHOCYTES | Scale | 9 | Lymphocytes | ANALYSIS | |
| LB_PNN | Scale | 10 | Neutrophiles | BIOLOGY | |
| LB_PRO_CT | Scale | 11 | Procalcitonine | BIOLOGY | |
| LB_S_CREATININ | Scale | 12 | Serum Creatinin | BIOLOGY | |
| LB_WBC | Scale | 13 | Leucocytes | RADIOLOGY | |
| MH_CRF | Nominal | 14 | Chronic renal failure | MEDICAL HISTORY | |
| MH_CVD | Nominal | 15 | Cardiovascular disease | MEDICAL HISTORY | FOR CLUSTERING |
| MH_DM | Nominal | 16 | Diabetes mellitus | MEDICAL HISTORY | |
| MH_HTN | Nominal | 17 | Hypertension | BASELINE | |
| MH_IS | Nominal | 18 | Immunosuppresion | MEDICAL HISTORY | |
| MH_LUNG_DISEASE_BINAR | Nominal | 19 | Lung Disease | BIOLOGY | |
| MH_SMOKER_BINARY | Nominal | 21 | Smoker | Medical History | |
| MH_SYMPTOMATIC | Nominal | 22 | Symptomatic C19 | Medical History | |
| MH_WEIGHT | Scale | 23 | Weight | MEDICAL HISTORY | |
| RX_CT_GGO_PCT_BASE | Scale | 24 | Ground glass estimation (%) @ Baseline CT | BIOLOGY | |
| RX_CT_LOBAR_CDX_BASE | Nominal | 25 | Lobar condensation @ Baseline CT | IDENTIFICATION | |
| RX_CT_PA_DIAMETER_BASI | Scale | 26 | Pulmonary artery diameter @ Baseline CT | RADIOLOGY | |
| SYMPTOME_DAY | Scale | 27 | Day symptoms started | BASELINE | |
| CT1_VALUE | Scale | 156 | 1st Ct value | BASELINE | |

| Variable | Position | Label | Domain | Measurement Level | |
|---|---|---|---|---|---|
| ATBG_BACTRIM | Nominal | 74 | Antibiogram-Bactrim | ANTIBIOGRAM | |
| ATBG_COLISTINE | Nominal | 77 | Antibiogram-colistine | ANTIBIOGRAM | |
| ATBG_MINOCYCLINE | Nominal | 76 | Antibiogram-Minocycline | ANTIBIOGRAM | |
| ATBG_QUINOLONE | Nominal | 75 | Antibiogram-quinolone | ANTIBIOGRAM | |
| TRT_TOCI_IL6 | Scale | 121 | IL6 assay results | BIOLOGY | |
| ICU_FILTER | Nominal | 86 | ICU admissions | ICU PARAMETERS | |
| FIO2_HIGH | Scale | 136 | Highest FiO² | ICU PARAMETERS | |
| OUT_HEMORRHAGE | Nominal | 53 | Hemorrhagic event | ICU PARAMETERS | |
| OUT_ET | Nominal | 55 | Intubation | ICU PARAMETERS | |
| INTUBATION_DAY | Scale | 103 | Intubation Day | ICU PARAMETERS | |
| EXTUBATION_DAY | Scale | 104 | Extubation Day | ICU PARAMETERS | |
| OUT_ICU | Nominal | 54 | ICU Transfer | ICU PARAMETERS | |
| PEEP_HIGH | Scale | 135 | Highest PEEP | ICU PARAMETERS | |
| ICU_DAY | Scale | 101 | ICU transfer Day | ICU PARAMETERS | |
| RX_CT_LOBAR_CDX_BASE | Nominal | 25 | Lobar condensation @ Baseline CT | IDENTIFICATION | |
| MH_IS_ADD | Binary | 154 | Immunosuppression | INFECTION | |
| INF_NOSOCOMIAL_BINARY | Nominal | 95 | Infection nosocomiale | INFECTION | |
| PATHOGEN_PULMONARY | Nominal | 70 | Pulmonary pathogen | INFECTION | |
| THOGEN_EXTRA_PULMONA | Nominal | 71 | Extrapulmonary pathogen | INFECTION | |
| MH_OLP | Nominal | 20 | Obstructive lung disease | MEDICAL HISTORY | |
| OUT_LOS | Scale | 127 | Length of Stay | OUTCOME | |
| OUT_HEMORRHAGE_DAY | Scale | 129 | Day of Hemorrhagic event | OUTCOME | |
| OUT_COMPOSITE | Nominal | 94 | Composite Outcome (ICU, Intubation, Death) | OUTCOME | |
| OUT_DEATH_BINARY | Nominal | 48 | All-cause mortality | OUTCOME | POST CLUSTERING |
| OUT_PCFS_M0_BOOLEAN | Nominal | 49 | PCFS M0 binary | OUTCOME | |
| OUT_PCFS_M2_BOOLEAN | Nominal | 50 | PCFS M2 binary | OUTCOME | |
| ICU_STAY | Scale | 102 | ICU length of Stay | OUTCOME | |
| OUT_DEATH | Nominal | 47 | Mortality | OUTCOME | |
| OUT_DISCHARGE_APP | Nominal | 56 | Discharge status | OUTCOME | |
| OUT_DISCHARGE_PCSF | Ordinal | 98 | PCFS @ discharge | OUTCOME | |
| OUT_MONTH2_APP | Nominal | 57 | 2-month post-discharge status | OUTCOME | |
| OUT_MONTH2_PCSF | Ordinal | 99 | PCFS @ 2-Month | OUTCOME | |
| OUT_NEWS2 | Scale | 126 | News 2 Score | OUTCOME | |
| OUT_TEE | Nominal | 52 | Thromboembolic event | OUTCOME | |
| OUT_TEE_DAY | Scale | 128 | Day of Thromboembolic event | OUTCOME | |
| SOFA | Scale | 140 | SOFA | OUTCOME | |
| APACHE | Scale | 141 | APACHE | OUTCOME | |
| MH_HF | Nominal | 157 | Heart failure | OUTCOME | |
| WHO_SCORE | Ordinal | 100 | WHO Score | OUTCOME | |
| DELTA_CT_DAYS | Scale | 130 | n (won't consider mean of velocity but ratio of m | RADIOLOGY | |
| DELTA_GGO_PCT | Scale | 132 | Variation of GGO (si >0 = bad) | RADIOLOGY | |
| DELTA_PA_DIAMETER | Scale | 131 | Variation of PA diameter (mm) (si >0 = bad) | RADIOLOGY | |
| RX_CT_PNEUMO_MED_ALL | Nominal | 87 | eumomediastinum (initial CT and/or Follow-up ( | RADIOLOGY | |
| RX_CT_LOBAR_CDX_1ST_FU | Nominal | 43 | Lobar condensation @ 1st Follow-up | RADIOLOGY | POST CLUSTERING |
| TRT_IVM_DAY | Scale | 109 | Ivermectine start day | TREATMENT | |
| TRT_HCQ_DAY | Scale | 105 | Hydroxychloroquine start day | TREATMENT | |
| OUT_DV | Nominal | 51 | Ventral Decubitus | TREATMENT | |
| HUMID | Nominal | 66 | Humid | TREATMENT | |
| TRT_AC | Nominal | 31 | Anticoagulants | TREATMENT | |
| TRT_AP | Nominal | 32 | Antiplatelets | TREATMENT | |
| TRT_ASA | Nominal | 30 | pirin (0 = no, 1 = ~81mg, 2 = ~162mg, 3 = ~324m | TREATMENT | |
| TRT_ATB_1 | Nominal | 36 | Penicillin | TREATMENT | |
| TRT_ATB_2 | Nominal | 37 | Cephalosporin | TREATMENT | |
| TRT_ATB_3 | Nominal | 38 | Carbapenem | TREATMENT | |
| TRT_ATB_4 | Nominal | 39 | Aminosid | TREATMENT | |
| TRT_ATB_5 | Nominal | 40 | Quinolone | TREATMENT | |
| TRT_ATB_6 | Nominal | 41 | Glycopeptide | TREATMENT | |
| TRT_ATB_7 | Nominal | 42 | Bactrim | TREATMENT | |
| TRT_ATB_ASSOCIATION | Nominal | 34 | Antibiotics association | TREATMENT | |
| TRT_ATB_DAY | Scale | 123 | Antibiotics start day | TREATMENT | |
| TRT_ATB_FAMILY | Nominal | 35 | Antibiotics class | TREATMENT | |
| TRT_ATB_SCOPE | Nominal | 33 | Antibiotics scope | TREATMENT | |
| TRT_AZT | Nominal | 59 | Azithromycine | TREATMENT | |
| TRT_AZT_DAY | Scale | 107 | Azithromycine start day | TREATMENT | |
| TRT_AZT_DURATION | Scale | 108 | Azithromycine duration | TREATMENT | POST CLUSTERING |
| TRT_BIOTHERAPY | Nominal | 88 | Biotherapy (Baricitinib or tocilizumab) | TREATMENT | |
| TRT_BARI | Nominal | 64 | Baricitinib | TREATMENT | |
| TRT_BARI_DAY | Scale | 118 | Baricitinib start day | TREATMENT | |
| TRT_BARI_DURATION | Scale | 119 | Baricitinib duration | TREATMENT | |
| TRT_TOCI | Nominal | 65 | Tocilizumab | TREATMENT | |
| TRT_TOCI_CURES | Nominal | 29 | Tocilizumab number of cures | TREATMENT | |
| TRT_TOCI_DAY | Scale | 120 | Tocilizumab start day | TREATMENT | |
| TRT_TOCI_DOSE | Scale | 122 | Tocilizumab dose | TREATMENT | |
| TRT_TOCI_TIMING | Nominal | 90 | Tocilizumab Timing (0 = day 1, 1 = day2+) | TREATMENT | |
| TRT_GC | Nominal | 63 | Glucocorticoids | TREATMENT | |
| TRT_GC_DAY | Scale | 115 | Corticoids start day | TREATMENT | |
| TRT_GC_DURATION | Scale | 117 | Corticoids duration | TREATMENT | |
| TRT_GC_MAX | Scale | 116 | Corticoids Maximal dose | TREATMENT | |
| TRT_HCQ | Nominal | 58 | Hydroxychloroquine | TREATMENT | |
| TRT_HCQ_DURATION | Scale | 106 | Hydroxychloroquine duration | TREATMENT | |
| TRT_IVM | Nominal | 60 | Ivermectine | TREATMENT | |
| TRT_IVM_DURATION | Scale | 110 | Ivermectine duration | TREATMENT | |
| TRT_L_R | Nominal | 61 | Lopinavir/Ritonavir | TREATMENT | |
| TRT_L_R_DAY | Scale | 111 | Lopinavir/ritonavir start day | TREATMENT | |
| TRT_L_R_DURATION | Scale | 112 | Lopinavir/ritonavir duration | TREATMENT | |
| TRT_RMS | Nominal | 62 | Remsidevir | TREATMENT | |
| TRT_RMS_DAY | Scale | 113 | Remsidivir start day | TREATMENT | |
| TRT_RMS_DURATION | Scale | 114 | Remsidivir duration | TREATMENT | |

surv3 = survival analysis where population is every ICU patient, deaths = dead, censored = survived ICU (i.e. discharged from ICU)

### Group 1

| ICU/PCT | >= 0.1 | <0.1 |
|---|---|---|
| 0 | 26 | 158 |
| 1 | 4 | 19 |

**207**

ICU 1/PCT >= 0.5 n/o

ICU 1/PCT < 0.5 n/o

### Group 2

| ICU/PCT | >= 0.1 | <0.1 |
|---|---|---|
| 0 | 32 | 64 |
| 1 | 19 | 22 |

**137**

ICU 1/PCT >= 0.5 n/o

ICU 1/PCT < 0.5 n/o

### Group 3

| ICU/PCT | >= 0.1 | <0.1 |
|---|---|---|
| 0 | 17 | 75 |
| 1 | 11 | 43 |

**146**

ICU 1/PCT >= 0.5 n/o

| ICU 1/PCT < 0.5 | coef | exp_coef | 2.5 | 97.5 | p_value |
|---|---|---|---|---|---|
| **TRT_GC_DURATION** | -5.2273521 | 0.005 | 0.000 | 0.240 | 7.04.E-03 |
| TRT_ATB_3_yes | 5.5046977 | 245.844 | 3.932 | 15370.350 | 9.08.E-03 |
| TRT_AZT_DAY | 4.7441118 | 114.906 | 3.138 | 4206.902 | 9.81.E-03 |
| TRT_TOCI_CURES_1 | -0.5276876 | 0.590 | 0.090 | 3.881 | 5.83.E-01 |
| TRT_TOCI_CURES_2 | 5.0569302 | 157.107 | 2.641 | 9347.530 | 1.53.E-02 |

surv2 = survival analysis where population is only the living, deaths = discharged, no censored

### Group 1

| ICU/PCT | >= 0.5 | <0.5 |
|---|---|---|
| 0 | 26 | 158 |
| 1 | 4 | 19 |
| | | 207 |

| ICU 0/PCT >= 0.5 | coef | exp_coef | 2.5 | 97.5 | p_value |
|---|---|---|---|---|---|
| TRT_GC_DURATION | -2.181689 | 0.113 | 0.034 | 0.370 | 3.14.E-04 |
| TRT_ATB_4_yes | -2.69712 | 0.067 | 0.006 | 0.708 | 2.46.E-02 |
| TRT_AZT_DAY | -1.77009 | 0.170 | 0.049 | 0.589 | 5.16.E-03 |
| TRT_ATB_7_yes | -22.559509 | 1.59415E-10 | 0 | Inf | 9.98.E-01 |

ICU 1/PCT >= 0.5

| ICU 0/PCT < 0.5 | coef | exp_coef | 2.5 | 97.5 | p_value |
|---|---|---|---|---|---|
| TRT_ATB_3_yes | -1.3393841 | 0.262 | 0.124 | 0.553 | 4.44.E-04 |
| TRT_TOCI_DOSE | 0.9014159 | 2.463 | 1.181 | 5.136 | 1.62.E-02 |

ICU 1/PCT < 0.5

### Group 2

| ICU/PCT | >= 0.5 | <0.5 |
|---|---|---|
| 0 | 32 | 64 |
| 1 | 19 | 22 |
| | | 137 |

| ICU 0/PCT >= 0.5 | coef | exp_coef | 2.5 | 97.5 | p_value |
|---|---|---|---|---|---|
| CL_O2_NEED_1 | 0.489 | 1.631 | 0.482 | 5.524 | 4.32.E-01 |
| CL_O2_NEED_2 | -3.599 | 0.027 | 0.003 | 0.285 | 2.60.E-03 |
| TRT_IVM_DURATION | 0.599 | 1.820 | 0.985 | 3.360 | 5.57.E-02 |

ICU 1/PCT >= 0.5

| ICU 0/PCT < 0.5 | coef | exp_coef | 2.5 | 97.5 | p_value |
|---|---|---|---|---|---|
| TRT_ATB_6_yes | -1.248932 | 0.287 | 0.102 | 0.807 | 1.79.E-02 |
| TRT_ATB_DAY | 0.5196893 | 1.682 | 1.090 | 2.595 | 1.89.E-02 |
| TRT_GC_DURATION | -0.4540597 | 0.635 | 0.428 | 0.942 | 2.38.E-02 |

ICU 1/PCT < 0.5

### Group 3

| ICU/PCT | >= 0.5 | <0.5 |
|---|---|---|
| 0 | 17 | 75 |
| 1 | 11 | 43 |
| | | 146 |

ICU 0/PCT >= 0.5

ICU 1/PCT >= 0.5

| ICU 0/PCT < 0.5 | coef | exp_coef | 2.5 | 97.5 | p_value |
|---|---|---|---|---|---|
| TRT_GC_DURATION | -0.6507376 | 0.522 | 0.348 | 0.783 | 1.68.E-03 |
| TRT_GC_MAX | -0.4088704 | 0.664 | 0.483 | 0.915 | 1.22.E-02 |
| OUT_DV_yes | 0.7418228 | 2.100 | 1.221 | 3.611 | 7.34.E-03 |
| TRT_DOXY_yes | 2.0819768 | 8.020 | 2.165 | 29.714 | 1.83.E-03 |

| ICU 1/PCT < 0.5 | coef | exp_coef | 2.5 | 97.5 | p_value |
|---|---|---|---|---|---|
| TRT_GC_DURATION | -1.202 | 0.300 | 0.160 | 0.563 | 1.75.E-04 |
| TRT_ATB_1_yes | -1.773 | 0.170 | 0.054 | 0.535 | 2.47.E-03 |
| TRT_TOCI_DAY | -1.397 | 0.247 | 0.090 | 0.682 | 6.94.E-03 |
| PATHOGEN_PULMONARY_BINARY_yes | -1.406 | 0.245 | 0.073 | 0.820 | 2.25.E-02 |

surv1 = survival analysis where population is everyone (deceased and living), deaths = cf. "formula"

### Group 1

| ICU/PCT | >= 0.5 | <0.5 |
|---|---|---|
| 0 | 26 | 158 |
| 1 | 4 | 19 |

207

ICU 0/PCT >= 0.5 n/o

ICU 1/PCT >= 0.5 n/o

ICU 0/PCT < 0.5 n/o

ICU 1/PCT < 0.5 n/o

### Group 2

| ICU/PCT | >= 0.5 | Column1 |
|---|---|---|
| 0 | 32 | 64 |
| 1 | 19 | 22 |

137

ICU 0/PCT >= 0.5 n/o

ICU 1/PCT >= 0.5 formula

| Surv(INTUBATION_DAY, OUT_ET) | coef | exp_coef | 2.5 | 97.5 | p_value |
|---|---|---|---|---|---|
| PATHOGEN_PULMONARY_BINARY_yes | 4.211 | 67.404 | 1.613 | 2816.268 | 2.70.E-02 |
| TRT_ATB_4_yes | -21.758 | 0.000 | 0.000 | Inf | 9.99.E-01 |
| TRT_GC_MAX | -2.218 | 0.109 | 0.006 | 1.847 | 1.25.E-01 |

ICU 0/PCT < 0.5 formula

| Surv(OUT_HEMORRHAGE_DAY, OUT_HEMORRHAGE) | coef | exp_coef | 2.5 | 97.5 | p_value |
|---|---|---|---|---|---|
| TRT_ATB_6_yes | 2.267 | 9.654 | 1.609 | 57.937 | 1.31.E-02 |

formula

| Surv(INTUBATION_DAY, OUT_ET) | coef | exp_coef | 2.5 | 97.5 | p_value |
|---|---|---|---|---|---|
| TRT_ATB_DAY | -1.072 | 0.342 | 0.143 | 0.822 | 1.64.E-02 |

ICU 1/PCT < 0.5 formula

| Surv(ICU_DAY, ICU_FILTER) | coef | exp_coef | 2.5 | 97.5 | p_value |
|---|---|---|---|---|---|
| TRT_TOCI_DAY | -1.411 | 0.244 | 0.075 | 0.793 | 1.90.E-02 |
| TRT_GC_DAY | -1.894 | 0.150 | 0.043 | 0.525 | 2.98.E-03 |
| TRT_ASA_1 | 1.038 | 2.823 | 0.648 | 12.297 | 1.67.E-01 |
| TRT_ASA_3 | 2.042 | 7.704 | 1.100 | 53.971 | 3.98.E-02 |
| TRT_HCQ_DURATION | -19.868 | 0 | 0.000 | 0.042 | 1.97.E-02 |

### Group 3

| ICU/PCT | >= 0.5 | Column1 |
|---|---|---|
| 0 | 17 | 75 |
| 1 | 11 | 43 |

146

ICU 0/PCT >= 0.5

ICU 1/PCT >= 0.5

ICU 0/PCT < 0.5 formula

| Surv(OUT_LOS, OUT_DEATH_BINARY) | coef | exp_coef | 2.5 | 97.5 | p_value |
|---|---|---|---|---|---|
| TRT_GC_DURATION | -6.890311 | 0.001 | 0.000 | 1.051 | 5.17.E-02 |
| TRT_AP_yes | 3.301662 | 27.158 | 1.914 | 385.400 | 1.47.E-02 |

ICU 1/PCT < 0.5 formula

| Surv(ICU_DAY, ICU_FILTER) | coef | exp_coef | 2.5 | 97.5 | p_value |
|---|---|---|---|---|---|
| TRT_TOCI_CURES_2 | -8.296 | 0.000 | 0.000 | 0.006 | 2.10.E-07 |
| TRT_TOCI_CURES_1 | -3.310 | 0.037 | 0.004 | 0.354 | 4.31.E-03 |
| TRT_GC_DAY | -1.724 | 0.178 | 0.074 | 0.427 | 1.10.E-04 |
| TRT_ATB_6_yes | -1.489 | 0.226 | 0.088 | 0.576 | 1.86.E-03 |
| TRT_ATB_DAY | -1.373 | 0.253 | 0.128 | 0.502 | 8.21.E-05 |
| TRT_BARI_DURATION | -1.308 | 0.270 | 0.092 | 0.797 | 1.77.E-02 |
| TRT_ATB_7_yes | -1.206 | 0.299 | 0.098 | 0.916 | 3.45.E-02 |
| TRT_RMS_DURATION | -1.136 | 0.321 | 0.095 | 1.087 | 6.79.E-02 |
| TRT_ATB_2_yes | **1.607** | 4.988 | 1.986 | 12.524 | 6.24.E-04 |
| PATHOGEN_PULMONARY_BINARY_yes | **3.176** | 23.947 | 7.322 | 78.321 | 1.50.E-07 |
| TRT_TOCI_TIMING_yes | **3.316** | 27.556 | 3.054 | 248.607 | 3.13.E-03 |

### Group 1

| ICU/PCT | >= 0.5 | <0.5 |
|---|---|---|
| 0 | 26 | 158 |
| 1 | 4 | 19 |
| | | 207 |

ICU 0/PCT >= 0.5
INF_NOSOCOMIAL_BINARY n/o
OUT_COMPOSITE n/o

ICU 1/PCT >= 0.5
INF_NOSOCOMIAL_BINARY n/o
OUT_COMPOSITE n/o

ICU 0/PCT < 0.5
INF_NOSOCOMIAL_BINARY n/o
OUT_COMPOSITE n/o

ICU 1/PCT < 0.5
INF_NOSOCOMIAL_BINARY n/o
OUT_COMPOSITE n/o

### Group 2

| ICU/PCT | >= 0.5 | <0.5 |
|---|---|---|
| 0 | 32 | 64 |
| 1 | 19 | 22 |
| | | 137 |

ICU 0/PCT >= 0.5
INF_NOSOCOMIAL_BINARY n/o
OUT_COMPOSITE n/o

ICU 1/PCT >= 0.5
INF_NOSOCOMIAL_BINARY n/o

| OUT_COMPOSITE | OR | 2.50% | 97.50% | pvalue |
|---|---|---|---|---|
| (Intercept) | 0.220 | 0.054 | 0.619 | 1.25.E-02 |
| TRT_ATB_4_yes | inf | 0.000 | NA | 9.95.E-01 |
| TRT_GC_DURATION | 0.174 | 0.015 | 1.027 | 1.18.E-01 |

ICU 0/PCT < 0.5
INF_NOSOCOMIAL_BINARY n/o

| OUT_COMPOSITE | OR | 2.50% | 97.50% | pvalue |
|---|---|---|---|---|
| (Intercept) | 0.207 | 0.078 | 0.465 | 4.43.E-04 |
| TRT_ATB_4_yes | inf | 0.000 | inf | 9.99.E-01 |
| TRT_AZT_DURATION | Inf | 0.000 | NA | 9.92.E-01 |
| TRT_ATB_2_yes | 8.056 | 1.571 | 49.226 | 1.49.E-02 |

ICU 1/PCT < 0.5
INF_NOSOCOMIAL_BINARY n/o
OUT_COMPOSITE n/o

### Group 3

| ICU/PCT | >= 0.5 | <0.5 |
|---|---|---|
| 0 | 17 | 75 |
| 1 | 11 | 43 |
| | | 146 |

ICU 0/PCT >= 0.5
INF_NOSOCOMIAL_BINARY n/o
OUT_COMPOSITE n/o

ICU 1/PCT >= 0.5
INF_NOSOCOMIAL_BINARY n/o
OUT_COMPOSITE n/o

ICU 0/PCT < 0.5
INF_NOSOCOMIAL_BINARY n/o
OUT_COMPOSITE n/o

ICU 1/PCT < 0.5
INF_NOSOCOMIAL_BINARY n/o
OUT_COMPOSITE n/o
