## Supplementary File 1 for "Machine Learning for COVID-19 Patient Management: Predictive Analytics and Decision Support"

1. Selection of clustering method

The distance-based method, K-prototypes (27), and the 2 model-based methods, KAMILA (12) and LCM (13), were used for data clustering. The 26 variables used by the algorithms for clustering were: leukocytes, neutrophils, lymphocytes, ferritin, lactate dehydrogenase (LDH), procalcitonin (PCT), creatinine (CRN), C-reactive protein (CRP), and D-dimer for laboratories; age, weight, sex; comorbidities including chronic renal failure, cardiovascular disease, hypertension, diabetes, pulmonary disease, smoking; immunocompromise (e.g., cancer, systemic disease), lung disease, smoking, level of oxygen requirement (0: no requirement, 1: < 4 L/min, 2: 4-8 L/min, 3: high flow); symptomatic COVID-19, day of symptom onset (day 0 is the day of hospitalization), first CT-PCR value; and radiological signs on baseline CT, including the percentage of ground glass, lobar condensation, and pulmonary artery diameter.

First, the appropriate number of clusters was determined using the silhouette score (based on Gower distance), which is a measure of an individual's similarity to his or her cluster relative to other clusters, and the Harrel concordance index (C-index), which quantifies the model's ability to predict an outcome (in this case, patient survival and discharge). Next, the results of each model were evaluated for clinical relevance by assessing the difference in survival and hospital stay between clusters and the distinguishing characteristics of each cluster. Radar plots and absolute standardized mean differences (ASMDs) between clusters were used to quantify the ability of variables to differentiate between clusters, which will be referred to as "signature variables" if this difference is ≥ 0.3.

2. Detailed review of regression analysis

Multivariate logistic and survival analyses (Cox proportional hazards model) were used to investigate the effect of treatments and the influence of patient risk factors on survival and the development of adverse events. We first scaled the continuous variables and then adjusted for their multicollinearity by calculating their variance inflation factor (VIF). Variables were then iteratively removed if their VIF was greater than 5. The selection of models was done progressively according to the Bayesian information criterion (BIC). If present, potential interactions were included in the models if their Wald test and likelihood ratio allowed it. For logistic regression, the goodness of fit of the resulting models was studied by plotting and analyzing the observed and fitted Pearson and deviance residuals. The influence of outliers was investigated by studying the Cook distance, and observations with >0.25 distance were removed from the data, and the model was refitted and re-evaluated with the new set of patients.

As with the Cox regression analysis, the proportionality of the risks was tested in the resulting model, and those that did not agree with this assumption were stratified into several other models to calculate consensus HRs for the remaining variables.

### 3. Results and selection of clustering algorithm

The first step is to cluster the data set and choose the best number of clusters. The optimal number of clusters should have the highest Silhouette score, associated with a high C-index and a low p-value on the log-rank test for both survival studies. For patient death, the highest curve has better survival, whereas, for hospital discharge, the lowest curve has the fastest discharge, making it better.

The highest Silhouette score was attributed to a k = 3 for the K-prototype method, having a value of 0.119, associated with a C-index of 0.654 and significant differences between the survival curves of each cluster (P = 0.001 and P < 0.001). LCM had a Silhouette of 0.087 for k = 2 and a C-index of 0.557 with good differences in survival analyses (P = 0.05 and P = < 0.001). Similar to KAMILA, k =3 yielded a Silhouette at 0.101 that was associated with the highest C-index of 0.656 among the other methods, with significant differences in survival studies for k =3 (P < 0.0001 and P < 0.0001). Figure 1 visualizes the silhouette plots for each number k of clusters for each method. To select the signature variables for each cluster, the ASMD was calculated for each pair of clusters in each method, and then the mean was deduced. Figure 2 shows the scores and discriminant values for each of the 26 features. The K-prototypes algorithm produced 15 signature features (Figure 2A), and 10 features for LCM (Figure 2C). However, KAMILA produced 19 signature features (Figure 2B).

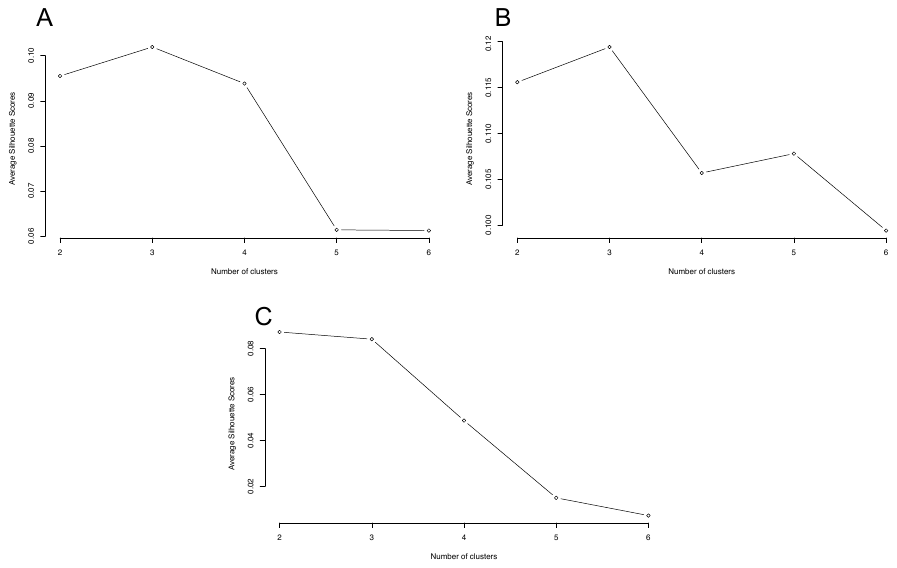

*Figure 1. Silhouette scores as a function of the number "k" of clusters in each algorithm. The k associated with the highest peak was chosen. A. KAMILA, B. K-prototypes, C. LCM*

To choose the best method, we decided to create an index that is nothing but the product of the Silhouette score, the C-index, and the number of signature features for each method. The method with the highest product will be elected as the method of choice. K-prototype had a product of 1.171, higher than LCM at 0.486. KAMILA had the highest product at 1.258, making KAMILA the method of choice, whose results will be used for the following statistical analyses.

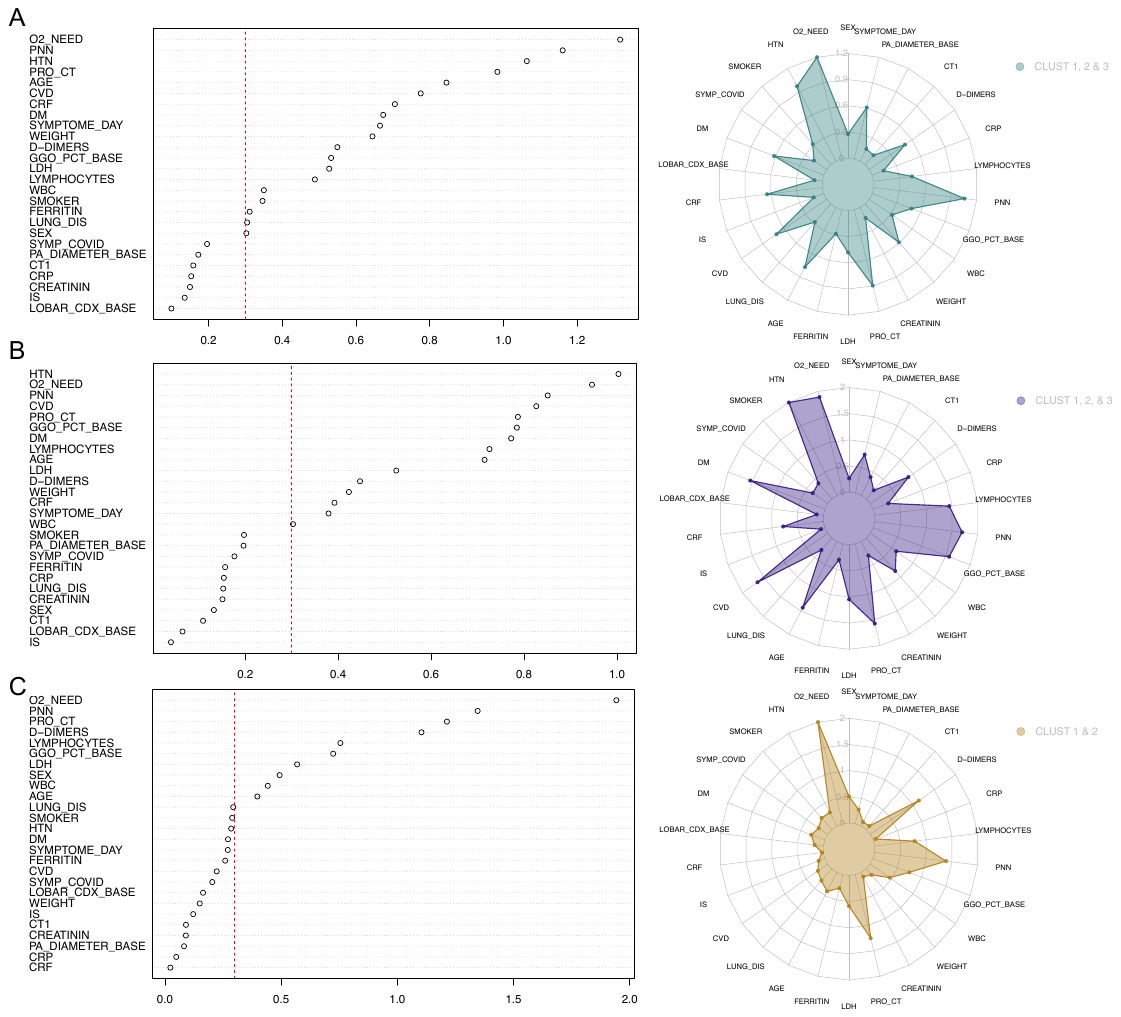

Figure 2. Average ASMD of the 26 variables in each algorithm. An ASMD >0.3 indicates that the variable should be considered to characterize clusters. A. KAMILA, B. K-prototypes, C. LCM

4. Variables used for regression analysis.

Oxygen supplementation; prone position; anticoagulation; antiplatelet therapy; aspirin therapy; date and duration of azithromycin therapy; date and duration of Baricitinib therapy; date, duration, dose, and number of tocilizumab courses; date, duration, and maximum glucocorticoid dose; date and duration of treatment with Lopinavir/Ritonavir; date and duration of treatment with Remsidevir; duration of treatment with Hydroxychloroquine; duration of treatment with Ivermectin; whether the patient was treated with Penicillin, Cephalosporin, Carbapenem, Aminoside, Doxycycline, Quinolone, Glycopeptide, and Bactrim; and the day of antibiotic initiation. In addition, and exclusively in this section of the analysis, we started the count from the day symptoms began; the count in the previous sections started at admission.

|  | Cluster 1 | Cluster 2 | Cluster 3 | All Clusters |
| --- | --- | --- | --- | --- |
| *Starting day of antibiotics* | 6.453 ± 5.62 | 5.555 ±5.82 | 9.392 ±5.68 | 7.038 ±5.885 |
| *Starting day of Azithromycin* | 5.095 ± 4.909 | 4.432 ±5.386 | 7.658 ±5.841 | 5.638 ±5.469 |
| *Duration of Azithromycin treatment* | 5.115 ±3.226 | 4.538 ±1.411 | 5.207 ±1.46 | 5.022 ±2.456 |
| *Starting day of Baricitinib* | 5.679 ± 5.103 | 4.961 ±5.869 | 9.899 ±6.66 | 6.678 ±6.138 |
| *Duration of Baricitinib treatment* | 9.667 ±4.243 | 10.778 ±5.63 | 9.643 ±5.982 | 9.969 ±5.301 |
| *Starting day of corticosteroids* | 5.695 ± 5.431 | 5.665 ±5.653 | 8.304 ±5.37 | 6.428 ±5.593 |
| *Duration of steroid therapy* | 20.144 ±19.026 | 21.208 ±17.086 | 24.7 ±18.101 | 22.011 ±18.222 |
| *Maximum dose of steroids* | 13.168 ±10.946 | 14.468 ±9.364 | 16.878 ±8.43 | 14.821 ±9.782 |
| *Duration of hydroxychloroquine therapy* | 9.492 ±4.232 | 12.762 ±20.231 | 10.5 ±4.554 | 10.345 ±9.374 |
| *Duration of ivermectin treatment* | 2.35 ±1.494 | 2.647 ±1.39 | 3 ±1.357 | 2.664 ±1.431 |
| *Day of initiation of Lopinavir/Ritonavir* | 7.133 ±2.924 | 8 ±2.828 | 7.5 ±3.536 | 6.441 ±5.634 |
| *Duration of Lopinavir/Ritonavir treatment* | 5.65 ± 5.003 | 4.871 ±5.868 | 9.196 ±5.367 | 7.263 ±2.806 |
| *Starting day of Remdesivir* | 0.733 ±0.884 | 2 ± 0 | 1.5 ±0.707 | 0.889 ±0.9 |
| *Duration of Remdesivir treatment* | 6.083 ±3.118 | 5 ±1.512 | 6.412 ±3.001 | 6 ±2.779 |
| *Starting day of Tocilizumab* | 5.819 ± 5.135 | 5.265 ±6.153 | 9.684 ±5.563 | 6.236 ±5.446 |
| *Dose of Tocilizumab* | 598 ±97 | 660 ±124 | 636 ±165 | 634 ±143 |

Table A - Mean values and standard deviations for regression analyses
